# CalCORVID: A Dynamic RShiny Dashboard Approach to Visualize Spatiotemporal Clusters for Public Health Surveillance

**DOI:** 10.1101/2025.08.29.25334603

**Authors:** Phoebe Lu, Seema Jain, Tomás M. León, Lauren A. White

## Abstract

**Background:** Infectious disease surveillance is an essential component of public health for preventing and mitigating outbreaks. Systematically applying statistical methods for anomaly detection to surveillance data can expedite outbreak response through early warning. A commonly used approach is the usage of spatiotemporal scan statistics as implemented in SaTScan, a software that analyzes spatiotemporal data to identify clusters of events over space and time that deviate from expected values. Some health departments identify outbreaks and prioritize resources using SaTScan for early cluster detection for diseases such as salmonellosis, legionellosis, and COVID-19. However, as a standalone software, SaTScan v10.2.1 does not provide functionality to easily disseminate visual cluster results over time in a way that is tailored to epidemiologists’ needs for real-time disease surveillance.

**Results:** We developed an open source dashboard that provides a customizable framework for displaying results and facilitating the use of SaTScan for public health surveillance. The California Clustering for Operational Real-time Visualization of Infectious Diseases (CalCORVID) dashboard is built using RShiny, is specifically designed for SaTScan outputs, and can be easily adapted to display any jurisdiction’s results. This dashboard features a map and corresponding results table, the option to view historical results, integration of the Social Vulnerability Index (SVI) to contextualize clusters, and interactive elements to enhance usability for epidemiologists.

**Conclusions:** We present CalCORVID as a complementary tool to native outputs of SaTScan v10.2.1, allowing users to visualize, customize, and distribute their results for specific public health use cases. Epidemiologists currently using SaTScan can adapt the provided code repository and dashboard template to display their own jurisdictions’ results, facilitating dissemination of cluster results for real-time, ongoing disease surveillance.

## Background

Infectious disease surveillance has many goals that include monitoring disease trends, establishing the current burden of disease, and identifying outbreaks (1). Responding to disease outbreaks is a critical responsibility of health departments to prevent further transmission and to find potential outbreak sources (2). To identify outbreaks, health departments typically rely on either facilities reporting outbreaks or epidemiologists monitoring the disease surveillance data to empirically determine whether there are anomalous patterns that merit investigation (2). Both approaches result in outbreaks that may have spread widely before they are detected and addressed; therefore, methods that can reduce the time to outbreak detection are of great interest to public health practitioners.

Spatiotemporal scan statistics, as implemented by SaTScan^TM^ software, are one of the methods used in applied public health settings to improve time to outbreak detection (3–5). In SaTScan, users provide an input dataset aggregated to a geographic unit (e.g., census tracts) along with the corresponding dates and associated number of cases or events. In applied public health settings, this input dataset often contains electronic laboratory reports because these are typically the first data source that a health department receives. The scan statistic conceptualizes each potential cluster as a 3-dimensional cylinder with the base and height corresponding to the spatial and temporal units, respectively. The scan statistic creates all possible combinations of these cylinders over a specified spatial area and temporal window, resulting in a large number of cylinders to scan over. Then, the scan statistic identifies anomalous clusters as cylinders with greater or lower observed events than expected when set to scan for high or low rates, respectively. For further technical details, please refer to Kulldorff et al. (6).

SaTScan has been adopted by several public health departments for routine surveillance. The New York City Department of Health and Mental Hygiene automates these spatiotemporal analyses for over 35 reportable diseases and has utilized results for early outbreak detection and resource allocation (3,7,8). Similarly, the New Jersey Department of Health implements SaTScan to aid in legionellosis surveillance and has used results for early outbreak detection and identification of additional cases (9). Other health departments have also successfully utilized the scan statistic to identify outbreaks (9–13).

Although there is existing documentation and literature to aid in the development of a SaTScan model (6,14), the dissemination of results for ongoing surveillance is less straightforward. Based on the selected output options, the SaTScan software produces several files for users to view results. The main output of SaTScan is a text file containing anomalous clusters listed in order of decreasing test statistic and associated cluster information. For users who want to manually visualize results, SaTScan provides spatial files in Keyhole Markup Language (KML) and shapefile formats that overlay the clusters on a map when imported into geographic software. Opening the KML file displays the clusters in Google Earth software while the shapefiles must be exported to geographic software (6). Both approaches require some manual configuration for the end user; the KML file must be in a shared file location that all users can access and have Google Earth software installed, and the shapefiles require an understanding of how to manipulate spatial data or the user of licensed spatial software. These requirements may exceed technical and resource capacity for some health departments and, in the case of ongoing monitoring, introduces repetitive, manual, non-reproducible steps into the surveillance process. The built-in visualization solution for SaTScan users is a Hypertext Markup Language (HTML) file that overlays the model results on a map with options to customize the view. Users can choose to display clusters by recurrence interval, change the map zoom level, and remove or add grid lines indicating the latitude and longitude coordinates. These native SaTScan outputs can be shared either through an automated email that is sent once the model analysis completes or by users accessing the files in a shared file location. However, maintaining a list of users for the automated email list or configuring file sharing permissions can be cumbersome or impractical, and users may also want to tailor results for different audiences (e.g., sharing line list information for internal audiences only). An open source, R-based solution to display and share results is a tractable alternative approach with the growing usage of R and RShiny in public health.

In California, several of our 61 local health jurisdictions have already implemented SaTScan in some form as part of their disease surveillance efforts. We developed California Clustering for Operational Real-time Visualization of Infectious Diseases (CalCORVID) in conversation with epidemiologists across California to address their need for an accessible solution to display and share their cluster results in real time. CalCORVID is an open source, interactive dashboard built in RShiny that facilitates visualization, customizability, and distribution of SaTScan results. While there are other SaTScan-related dashboards, these dashboards are wrappers for the rsatscan package that enable running SaTScan within the R software (15,16). Our open source SaTScan dashboard is distinguished by its focus on use in applied public health settings and includes features such as the ability to customize the results display, view clusters over time, interact directly with cluster results, and incorporate socioeconomic spatial variables to contextualize findings.

## Implementation

Key features of the CalCORVID dashboard include:

- Displaying cluster results without requiring any manipulation of spatial objects, which improves dashboard processing and usability
- Results tab that features a map displaying clusters and corresponding cluster information
- Date drop-down element to view historical data and compare results visually over time
- Ability to visualize clusters overlaid with Social Vulnerability Index (SVI) to help contextualize results
- Reactive elements to interact with SaTScan cluster results
- Technical notes tab to describe model details for end user
- Option to automate ongoing surveillance tasks using RShiny framework

## Implementation Details

CalCORVID is written in the R programming language and is released under the MIT Open Source License (https://opensource.org/license/mit/). The current distribution includes CalCORVID dashboard source code, tutorials on how to use the tool, and sample datasets. The documentation also describes the overall structure of the dashboard and basic RShiny functionalities for individuals who may be interested in customizing CalCORVID for their own use.

The primary output of this repository is an open source dashboard code for users to adapt for their spatiotemporal cluster outputs from SaTScan. This dashboard can be used by epidemiologists conducting routine surveillance or sharing with other stakeholders who may be interested in visualizing disease clusters. The documentation describes the input files needed to display clusters correctly on the dashboard and the parameters to change for successful adaptation to individual use cases.

CalCORVID is distributed with example data from the California Open Data Portal to demonstrate data preprocessing and dashboard display. An additional simulated dataset is provided and detailed in the README file to help users become more familiar with the structure of the dashboard before incorporating their own data.

### User interface and distribution

We developed the CalCORVID dashboard using R (version 4.0.4.) (17) and Shiny (version 1.7.1) (18). The required software and R packages necessary to run this dashboard are listed in Supplementary Table 1. By using the Shiny package, the dashboard can utilize reactive functions which allows the user to interact with widgets in the dashboard to filter and display further information.

The CalCORVID dashboard is based on two files that must be in the same directory: app.R and global.R. The app.R file contains code for the user interface (UI) and the server functions. Elements displayed on the dashboard’s visual interface are stored in the UI object, and must first be rendered by the server functions. The global.R file must be sourced prior to running app.R, which loads libraries, preprocesses data, loads data, and allows the user to specify function options. Although the global.R file is not required by Shiny, loading and preprocessing data in this file improves the performance of app.R.

In addition to these three files, the repository contains data, R and www.folders, which are essential to run this dashboard. The data folder contains all the relevant raw data files needed for the dashboard, with subfolders for different inputs such as the coordinates file (“coords”), county boundary shapefiles (“county_boundary”), combined “gis” and “col” files (“giscol”), SaTScan cluster outputs (“satscan_output”), and Social Vulnerability Index (SVI) files (“svi”). The R folder holds the R scripts containing functions that are used to generate the data in the global.R file. Lastly, the www.folder is RShiny-specific and is the directory that locally stores elements that are rendered in the web browser (e.g., images for the home page).

### Data generating process and input file

Generating the dashboard input dataset is a separate process from what this open source repository provides. As part of their own workflow, users must obtain comma-separated value (CSV) files of cluster results after running their data in the SaTScan software, which can be specified from the SaTScan Outputs tab and checking the boxes corresponding to “Cluster Information” and “Location Information.” Checking these boxes will output files with “col” and “gis” file extensions, respectively. The “col” file contains the cluster centers and the columns the dashboard requires as shown in Table 1. The “gis” file contains the associated location IDs with each cluster center, which are used to calculate location-level spatial covariates.

**Table 1.** Columns required for raw input data for both .col and .gis files.

| SaTScan Variable | Description | Originating File |
| --- | --- | --- |
| CLUSTER | Assigned cluster number. This number is assigned by descending log likelihood ratios (i.e., the first cluster will have the highest log likelihood value.) | .col, .gis |
| LOC_ID | Cluster center (.col) or location ID (.gis) in the given geographic unit .<br>For example, if analysis is at the census tract level, these will correspond to the census tract at the center of the cluster and census tracts associated with each cluster, respectively. | .col, .gis |
| LATITUDE | Cluster center latitude coordinate. | .col |
| LONGITUDE | Cluster center longitude coordinate. | .col |
| RADIUS | Cluster center radius in kilometers. | .col |
| START_DATE | Cluster start date. | .col |
| END_DATE | Cluster end date. This date will always correspond to the most recent date your dataset is run on. For example, if the last date of your analysis is 1/30/24, the clusters will all have the end date of 1/30/24. | .col |
| OBSERVED | Observed number of cases in cluster between START_DATE, END_DATE, and associated LOC_IDs. | .col |
| EXPECTED | Expected number of cases in cluster between START_DATE, END_DATE, | .col |
|  | and associated LOC_IDs. |  |
| RECURR_INT | Recurrence interval, or how often the observed cluster would be observed by chance, assuming analyses are repeated on a regular basis with a periodicity equal to the specified time interval length (6). For example, a recurrence interval of 100 days indicates that a cluster like the observed cluster would be observed only every 100 days (occurrence is unusual.) | .col |

For demonstration purposes, we include a sample results file generated from California COVID-19 vaccination data. We used publicly available, weekly California vaccination data aggregated at the ZIP code level from the California Open Data Portal, sourced June 3, 2024 (19). We ran a prospective space-time analysis with the discrete Poisson probability model to detect low clusters of vaccination events for an arbitrary time frame from September 21, 2021, to February 8, 2022, to provide sample SaTScan results for display in the dashboard (20). Supplementary Table 2 details the specific model parameters used to generate the sample results.

### Pre-processing files

After the user obtains the required output files, some data preprocessing is necessary before running the dashboard. Table 2 details the different functions provided with the open source repository that facilitate this process, and sample code is provided in the global.R file. These functions are sourced and automatically run by global.R when app.R is run, but users can opt to run these functions individually to debug their code.

**Table 2.**
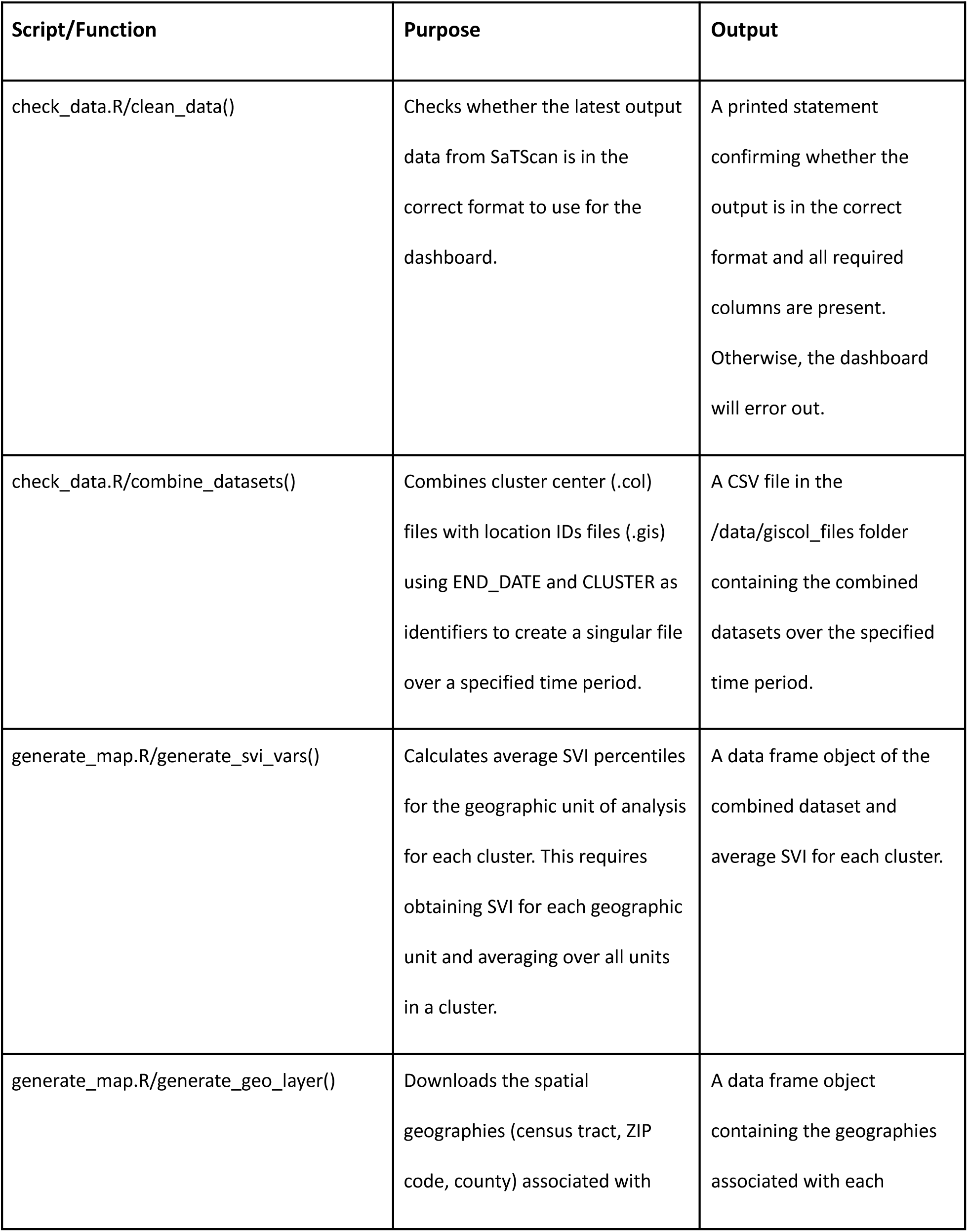
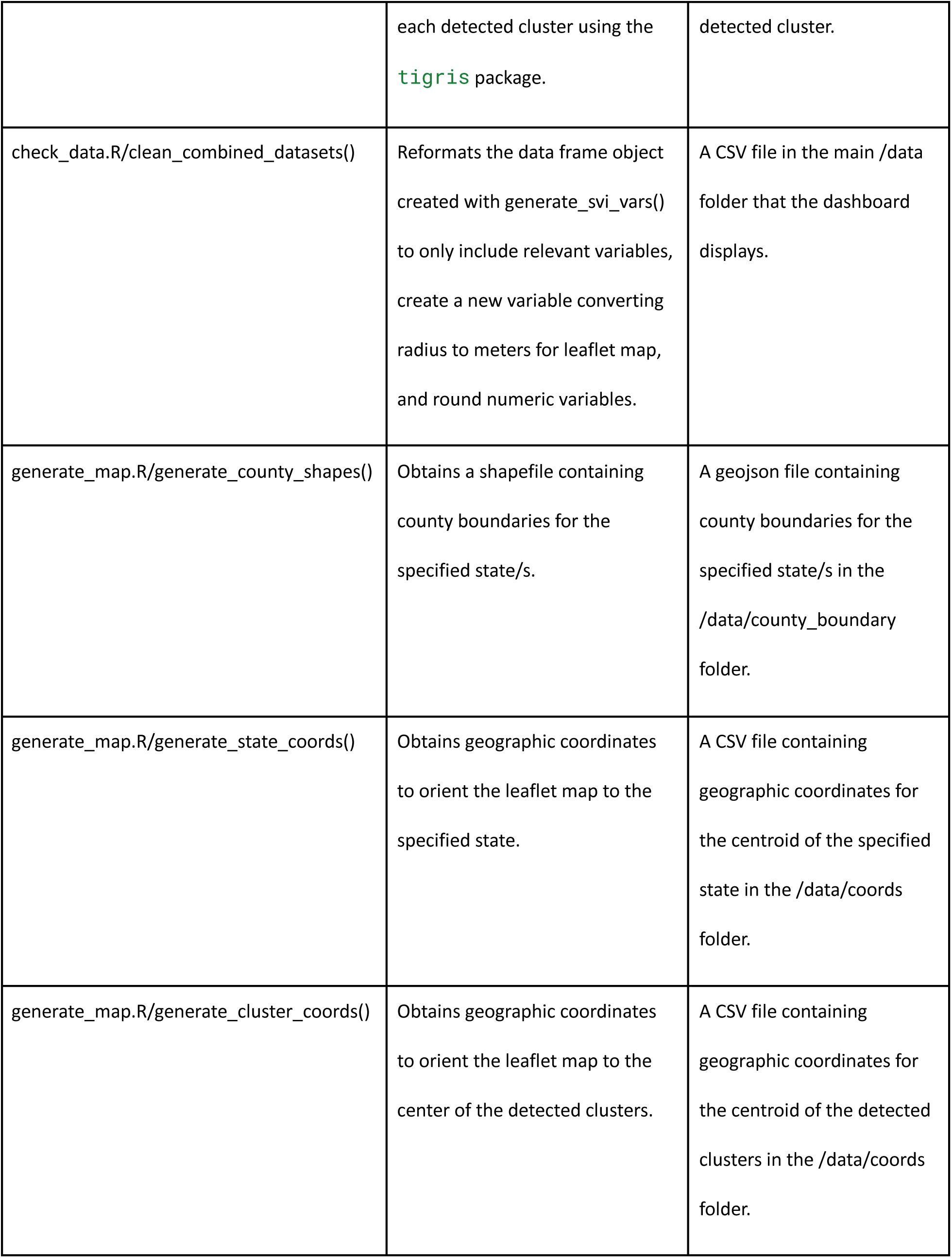
Functions included in the dashboard in the R folder, listed in the necessary order for preprocessing.

Briefly, global.R first checks whether their latest output files are in the correct formats for the dashboard using the clean_data() function. If they are, the historical file can be created with the combine_datasets() function which combines the cluster center and location ID files over the specified historical period. After combining the datasets for the desired time period, the average Social Vulnerability Index (SVI) percentiles across all detected geographies for each cluster are calculated using the generate_svi_vars() function. SVI is a socioeconomic index created by the CDC based on 16 US census variables and is commonly used to compare the social vulnerability between different geographic units (21). There are four themes with percentile rankings: socioeconomic status, household characteristics, racial and minority status, and housing type and transportation, and an overall ranking for vulnerability (22). Since the percentile ranking for a geographic unit is calculated relative to other geographic units in the dataset, percentiles may be slightly different depending on the unit of analysis chosen; therefore, the average percentile for a cluster containing ZIP code units will differ from the average percentile for a cluster containing census tract units, even if the ZIP codes contain all census tracts. After the SVI metrics are calculated for each cluster, we use generate_geo_layer() to obtain the spatial geometries for each of the detected clusters for display on the leaflet map. The last step is the clean_combined_datasets() function that reformats the combined file containing average SVI percentiles and spatial geometries for display on the CalCORVID dashboard. This generates a CSV file in the /data/ folder on which the dashboard is based.

There are also two additional functions within global.R that are run the first time a new analysis is displayed on the dashboard. The generate_county_shapes() function creates a geojson spatial file that contains the county boundaries for the state or states encompassing the data. The resulting object is used as a layer with Leaflet, an open source JavaScript library used for interactive mapping, and will be detailed in a later section (23). The R leaflet package adapts Leaflet functionality into R to generate the main basemap display on the CalCORVID dashboard (24). After the base Leaflet map is created, we overlay spatial files such as county boundaries to provide additional spatial information. The second function that runs is either the generate_state_coords() or generate_cluster_coords() function, depending on the zoom level specified in the parameter settings. The generate_state_coords() function orients the Leaflet map to zoom to the state level of the data and the generate_cluster_coords() function orients the map to the center of the detected clusters.

## Dashboard Features

### Sample dataset and date dropdown menu

The dataset generated by the clean_combined_datasets() function is the underlying dataset for the CalCORVID dashboard. Our sample final dataset is named CAvax_combgiscol_fnl.csv and is located in the /data/ folder. The main reactive functions filtering the map and table are the date dropdown and recurrence interval slider elements, which allow users to filter display results by date or recurrence interval, respectively. The available date range is determined in the combine_datasets() preprocessing step described above.

### Leaflet map displaying visual clusters

The map is the main visual element of the results page. First, we render the map with our selected base map using renderLeaflet() and set the default view to the state of analysis. The geographic coordinates used to set the view are generated either by the generate_state_coords() or generate_cluster_coords() functions provided in the R folder depending on the parameter specified in the global.R file. After rendering the map, we use leafletProxy() to add features to the map. Using leafletProxy() improves the processing time of the dashboard because it does not need to render the base map every time the dashboard is reloaded. We use leaflet::addPolygons() to display the location geographies associated with each detected cluster, randomly assigning colors to more easily distinguish clusters. Clicking on these clusters results in a pop-up containing information about the average SVI for each cluster. For circular scans we provide an optional overlay of the circles using leaflet::addCircles(), which draws the circles using the geographic coordinates of the cluster center and radius in meters. This approach allows for the display of additional information without compromising dashboard loading times by only requiring a CSV file with the coordinates and radii instead of loading multiple shapefiles. We also use the leaflet::addLayersControl() function to create a toggleable layer for displaying county boundaries, which are generated using the generate_county_shapes() function provided in the R folder.

### Data table displaying cluster information

Building the table requires converting the dataset to a reactive element using the reactive() function so reactive elements such as the date dropdown will work. Then, we convert the reactive dataset to a datatable widget using the datatable() function from the DT package, select the datatable display options, select variables of interest to display, and rename variables to be user-friendly. The display options for the datatable object can be found in the documentation for the DT package (25). Finally, we render the table using the renderDT() function, which allows the table to be displayed in the user interface using the DTOutput() function.

In addition to displaying the cluster data, clicking a row in the table drops a pin on the corresponding cluster on the map to make it easier to identify. This reactive element uses the observeEvent() function to identify the row that is clicked and highlight the cluster it belongs to. Clicking on the row again clears the pin.

## Results/Discussion

### Dashboard

The final CalCORVID dashboard consists of a homepage, a main results page, and a technical notes page. The homepage serves as the landing page for users when they first load the dashboard with the option for users to navigate onward to the main results page (Figure 1A) or technical notes page. The homepage is customizable and can be configured with different icons and personalized with the jurisdiction’s contact information at the bottom if shared with stakeholders. The main results page contains the date dropdown, map, and table elements described in the Implementation section. The date dropdown allows the user to select results from a predefined list of dates generated during the data preprocessing step using the combine_datasets() function. This functionality helps epidemiologists visually track whether clusters become more or less statistically significant over time and whether they merit investigation. The main results page contains several other features that facilitate usability and provide further epidemiologic context (Figures 1A-C). Clicking on a row in the cluster results table highlights the corresponding cluster in the map as shown (Figure 1A). This interactivity ties the two visualizations together and aids in identification of map clusters, which can be difficult when multiple clusters are displayed. When clicking on a cluster on the map, the tooltip displays additional information about spatial covariates incorporated into the map (Figure 1B). In this use case, the tooltip contains the average overall SVI and average SVI for each of the four SVI themes over all the geographic units included in the cluster. Finally, users have the option to select and view the county boundary layer generated by the generate_county_shapes() function which is used to geographically orient the user (Figure 1C).

**Figure 1.**
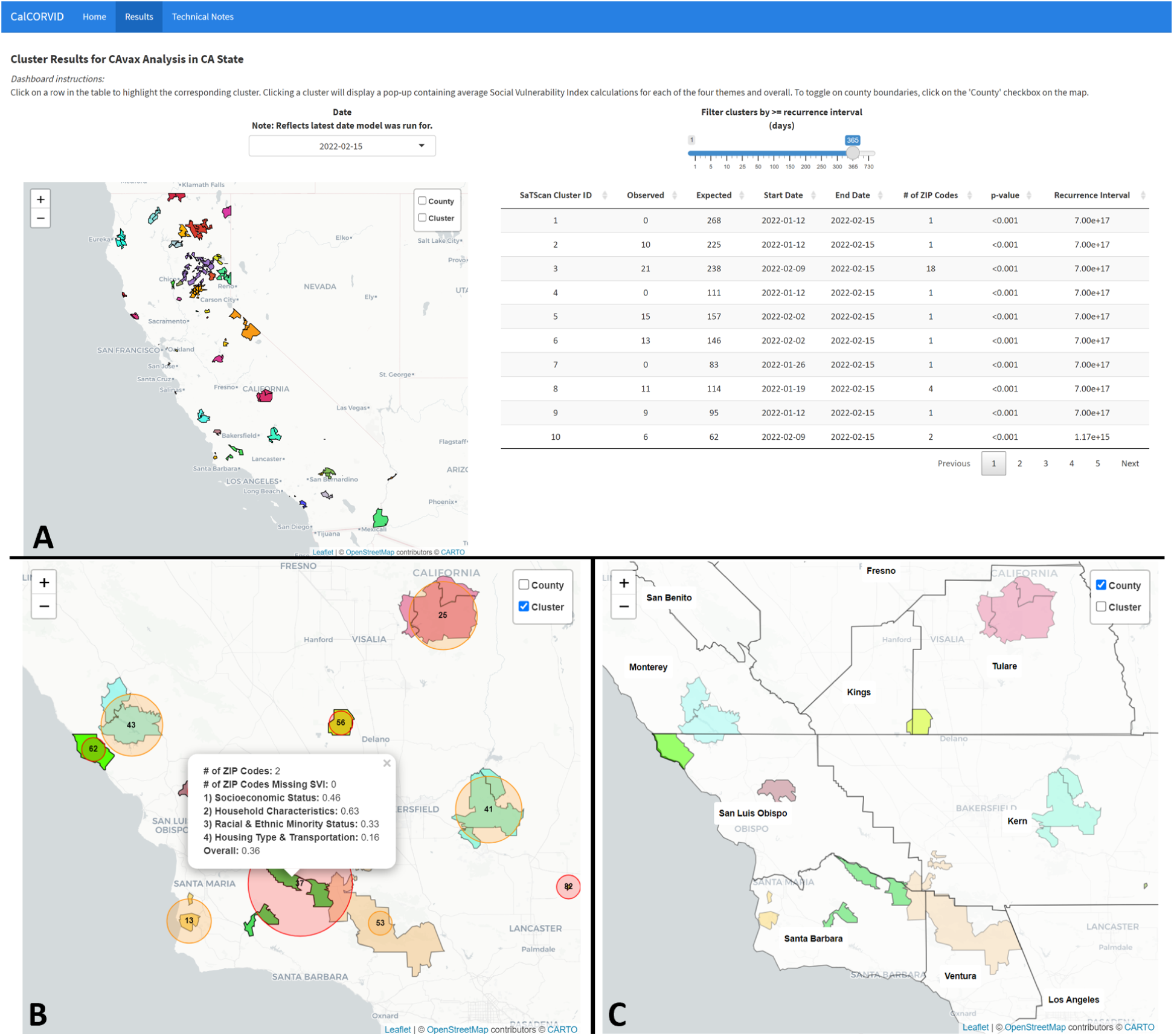
(A) Main results page with the analysis name and state of analysis defined at the top of the page. A date drop down above the map allows users to switch between different dates for map and table results and a slider is provided to filter by recurrence interval. The map is set to the “state” zoom level. Clicking on a table row will highlight the corresponding cluster on the map to identify clusters more easily, with each cluster distinguished by a randomly selected color. (B) Checking the “Cluster” layer overlays the circular cluster scan on the cluster boundaries. Clicking on a cluster on the map provides the average Social Vulnerability Index (SVI) metrics for each of the four themes, the overall SVI, and the number of geographic units in the cluster missing SVI data. (C) Checking the “County” layer on the map displays the county boundary layers to help spatially orient the user.

### California Department of Public Health Workflow and Implementation

We describe our current implementation to provide an example of how other jurisdictions may want to integrate the dashboard into their own processes. The California Department of Public Health (CDPH) is interested in methods for identifying spatiotemporal clusters of disease to understand trends across the state. We run an automated prospective space-time analysis with a discrete Poisson probability model using nucleic acid amplification test (NAAT) results as our input data (7). This allows us to monitor COVID-19 test positivity trends state-wide and regionally. Our data workflow is broken down into five distinct steps as shown in Figure 2. We pull NAAT testing data from our data warehouse weekly and preprocess data in R to aggregate individuals to the census tract level and format the input files for SaTScan. Once the input files are created, we call the SaTScan software within R using the rsatscan package and run our analyses. After obtaining our results, we conduct a bipartite network analysis on the clusters to analyze whether they are part of the same extended cluster. Although we do not detail this process in the paper, this step is intended to distinguish unique spatial patterns over multiple overlapping clusters. Finally, the processed cluster results are output to the RShiny dashboard for local health departments in California to access. We automate our pipeline using an online server and schedule each job to run sequentially. The dashboard automatically updates with new data. Local health departments and epidemiologists use this dashboard as a situational awareness tool to understand COVID-19 trends in their region and at the state-wide level.

**Figure 2.**
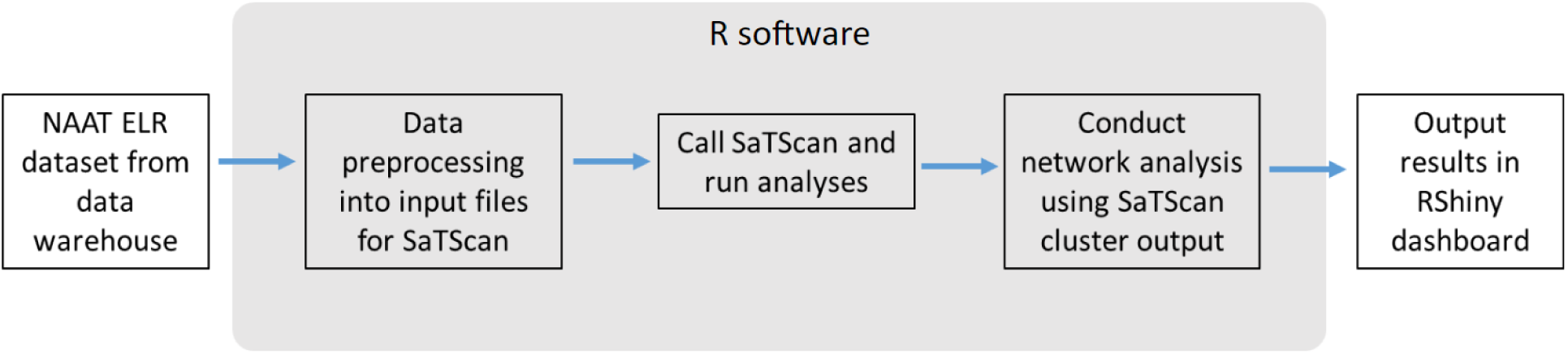
Flow diagram of CDPH workflow culminating in the RShiny dashboard display of results. We pull our NAAT electronic laboratory reports (ELRs) from our data warehouse, preprocess the testing data into input files for SaTScan, run the SaTScan software, apply a bipartite network analysis to the SaTScan cluster results, and output the final results into the CalCORVID-based dashboard.

### Comparison with Other Dashboards

Although other SaTScan-related dashboards exist, CalCORVID is the only one that explicitly addresses the challenges of visualizing spatiotemporal cluster results in an applied public health context. The most similar dashboard to CalCORVID is SpatialEpiApp, a RShiny-based dashboard that integrates SaTScan for cluster detection and disease risk estimation using integrated nested Laplace approximation (INLA) (16). The input page requires two files: a spatial file containing the underlying base map and a CSV file for the SaTScan input case file. The user can then choose from spatial or spatiotemporal analyses to run. The results page contains several outputs, including an interactive map, model results, cluster results, and a downloadable report. One of the main differences between SpatialEpiApp and CalCORVID is that SpatialEpiApp requires users to run their data through the dashboard’s SaTScan mechanism to display the visualizations, while CalCORVID does not. Another SaTScan-related dashboard is EpiExploreR, which is a dashboard that provides tools to analyze spatiotemporal data, explore input data, and visualize results (15). EpiExploreR combines multiple R packages and is distinguished by its ability to analyze epidemiological data using multiple methods, including SaTScan. Similar to SpatialEpiApp, EpiExploreR requires the user to run their data using the dashboard’s wrapper for SaTScan before results are displayed. Although both interfaces simplify running SaTScan for the user, the limited SaTScan parameter options may restrict users from more complex or nuanced analyses. For example, the SaTScan software allows users to select prospective or retrospective analyses, choose from eight different probability models, and scan for clusters with high and/or low rates while SpatialEpiApp only allows for prospective analyses, two probability models, and scans for high rates. SaTScan software also contains advanced options to refine cluster results, such as changing the spatial scan radius, temporal scan window, minimum number of cases to consider a cluster, and the ability to scan for clusters along a specified network. These advanced options are essential to correctly configure the spatiotemporal scan statistic for the disease of interest and jurisdiction (26). CalCORVID simply requires the SaTScan output files and can be adapted for a variety of different models or use cases. The CalCORVID dashboard also streamlines the results page so epidemiologists can quickly browse through results as part of their surveillance routine.

### Future Directions

The first iteration of CalCORVID is a RShiny dashboard that is easily adaptable for different SaTScan models and provides basic functions that facilitate epidemiologic surveillance activities. Future extensions of the dashboard include support for a quantitative method to track clusters over time, inclusion of additional socioeconomic variables beyond the Social Vulnerability Index, support for display of linelists associated with each cluster, and extension to other spatiotemporal cluster detection methods. We plan to maintain CalCORVID as an open source repository and welcome contributions and suggestions to improve the functionality of the dashboard.

## Conclusion

Spatiotemporal scan statistics are a useful complementary tool to traditional infectious disease surveillance activities. A dashboard is an effective method to visualize, monitor, and disseminate cluster results to a wide audience, and the open source code provided in the CalCORVID repository provides a starting point for epidemiologists interested in using the scan statistic as part of their workflow.

## Availability and Requirements

**Project name:** CalCORVID: A Dynamic RShiny Dashboard Approach to Visualize Spatiotemporal Clusters for Public Health Surveillance

**Project home page:** https://github.com/cdphmodeling/CalCORVID

**Operating system(s):** Windows, MacOS, Linux

**Programming language:** R 4.0 or higher

**Other requirements:** See Supplementary Table 1

**License:** MIT

**Any restrictions to use by non-academics:** N/A

## Data Availability

All data and code produced in the present work are contained in the following GitHub repository: https://github.com/cdphmodeling/CalCORVID

https://data.chhs.ca.gov/dataset/covid-19-vaccine-progress-dashboard-data-by-zip-code

## List of abbreviations

CalCORVID: California Clustering for Operational Real-time Visualization of Infectious Diseases
CDPH: California Department of Public Health
ELR: electronic laboratory report
SaTScan: Spatial and Temporal Scan (software)
SVI: Social Vulnerability Index

## Declarations

## Acknowledgements

The authors thank the members of the CDPH Modeling and Advanced Analytics Team including Héctor Manuel Sánchez Castellanos, Mugdha Thakur, Natalie Linton, and Sindhu Ravuri for conversations, insights, and suggestions that improved the code and implementation. We also thank Sharon Greene and Alison Levin-Rector from the New York Department of Health and Mental Hygiene for contributing invaluable feedback for the manuscript draft.

## Authors’ Contributions

PL designed the dashboard and wrote the manuscript. LW and TL revised and edited the manuscript. SJ supervised the project. All authors read and approved the final manuscript.

## Availability of data and materials

All data and code used to generate the dashboard is available in the public repository: https://github.com/cdphmodeling/CalCORVID

## Declarations

### Ethics approval and consent to participate

Not applicable.

### Consent for publication

Not applicable.

## References

1. Murray J, Cohen AL. Infectious Disease Surveillance. Int Encycl Public Health. 2017;222–9.

2. Reingold A. Outbreak Investigations—A Perspective. Emerg Infect Dis. 1998 Mar;4(1):21–7.

3. Greene S, Peterson E, Kapell D, Fine A, Kulldorff M. Daily Reportable Disease Spatiotemporal Cluster Detection, New York City, New York, USA, 2014–2015. Emerg Infect Dis. 2016 Oct 1;22.

4. Mostashari F, Kulldorff M, Hartman JJ, Miller JR, Kulasekera V. Dead Bird Clusters as an Early Warning System for West Nile Virus Activity. Emerg Infect Dis. 2003 Jun;9(6):641–6.

5. Viñas MR, Tuduri E, Galar A, Yih K, Pichel M, Stelling J, et al. Laboratory-Based Prospective Surveillance for Community Outbreaks of Shigella spp. in Argentina. PLoS Negl Trop Dis. 2013 Dec 12;7(12):e2521.

6. Kulldorff M. SaTScan User Guide v10.1 [Internet]. 2022 [cited 2024 Feb 13]. Available from: https://www.satscan.org/cgi-bin/satscan/register.pl/SaTScan_Users_Guide.pdf?todo=process_userguide_download

7. Greene SK, Peterson ER, Balan D, Jones L, Culp GM, Fine AD, et al. Detecting COVID-19 Clusters at High Spatiotemporal Resolution, New York City, New York, USA, June-July 2020. Emerg Infect Dis. 2021 May;27(5):1500–4.

8. Latash J. Salmonellosis Outbreak Detected by Automated Spatiotemporal Analysis — New York City, May–June 2019. MMWR Morb Mortal Wkly Rep [Internet]. 2020 [cited 2024 Jan 28];69. Available from: https://www.cdc.gov/mmwr/volumes/69/wr/mm6926a2.htm

9. Gleason JA, Ross KM. Development and Evaluation of Statewide Prospective Spatiotemporal Legionellosis Cluster Surveillance, New Jersey, USA. Emerg Infect Dis. 2022 Mar;28(3):625–30.

10. Jones RC, Liberatore M, Fernandez JR, Gerber SI. Use of a Prospective Space-Time Scan Statistic to Prioritize Shigellosis Case Investigations in an Urban Jurisdiction. Public Health Rep. 2006;121(2):133–9.

11. Sharip A, Bs JM, Croker C, Kim M, Hwang B, Aller R, et al. Preliminary Analysis of SaTScan’s Effectiveness to Detect Known Disease Outbreaks Using Emergency Department Syndromic Data in Los Angeles County.

12. Verma A, Schwartzman K, Behr MA, Zwerling A, Allard R, Rochefort CM, et al. Accuracy of prospective space–time surveillance in detecting tuberculosis transmission. Spat Spatio-Temporal Epidemiol. 2014 Apr 1;8:47–54.

13. Stelling J, Read JS, Peters R, Clark A, Bokhari M, O’Brien TF. Staphylococcus aureus antimicrobial susceptibility trends and cluster detection in Vermont: 2012-2018. Expert Rev Anti Infect Ther. 2021 Jun 3;19(6):777–85.

14. Levin-Rector A, Kulldorff M, Peterson E, Hostovich S, Greene S. Prospective Spatiotemporal Cluster Detection using SaTScan: A Tutorial for Designing and Finetuning a System to Detect Reportable Communicable Disease Outbreaks (Preprint). 2023.

15. Savini L, Candeloro L, Perticara S, Conte A. EpiExploreR: A Shiny Web Application for the Analysis of Animal Disease Data. Microorganisms. 2019 Dec;7(12):680.

16. Moraga P. SpatialEpiApp : A Shiny web application for the analysis of spatial and spatio-temporal disease data. Spat Spatio-Temporal Epidemiol. 2017 Nov;23:47–57.

17. R: The R Project for Statistical Computing [Internet]. [cited 2024 Jan 22]. Available from: https://www.r-project.org/

18. Chang W, Cheng J, Allaire JJ, Sievert C, Schloerke B, Xie Y, et al. shiny: Web Application Framework for R [Internet]. 2023 [cited 2024 Jan 22]. Available from: https://cran.r-project.org/web/packages/shiny/index.html

19. COVID-19 Vaccine Progress Dashboard Data by ZIP Code - California Health and Human Services Open Data Portal [Internet]. [cited 2024 Jun 3]. Available from: https://data.chhs.ca.gov/dataset/covid-19-vaccine-progress-dashboard-data-by-zip-code

20. Kulldorff M. A spatial scan statistic. Commun Stat - Theory Methods. 1997 Jan 1;26(6):1481–96.

21. CDC/ATSDR Social Vulnerability Index (SVI) [Internet]. 2024 [cited 2024 Jan 30]. Available from: https://www.atsdr.cdc.gov/placeandhealth/svi/index.html

22. CDC/ATSDR SVI Frequently Asked Questions (FAQ) | Place and Health | ATSDR [Internet]. 2022 [cited 2024 Jan 30]. Available from: https://www.atsdr.cdc.gov/placeandhealth/svi/faq_svi.html

23. Agafonkin V. Leaflet, a JavaScript library for interactive maps [Internet]. 2024 [cited 2024 May 17]. Available from: https://github.com/Leaflet/Leaflet

24. Cheng J, Schloerke B, Karambelkar B, Xie Y, Wickham H, Russell K, et al. leaflet: Create Interactive Web Maps with the JavaScript “Leaflet” Library [Internet]. 2023 [cited 2024 Jan 22]. Available from: https://cran.r-project.org/web/packages/leaflet/index.html

25. Using DT in Shiny [Internet]. [cited 2024 Feb 1]. Available from: https://rstudio.github.io/DT/shiny.html

26. Levin-Rector A, Kulldorff M, Peterson ER, Hostovich S, Greene SK. Prospective Spatiotemporal Cluster Detection Using SaTScan: Tutorial for Designing and Fine-Tuning a System to Detect Reportable Communicable Disease Outbreaks. JMIR Public Health Surveill. 2024 Jun 11;10:e50653.

27. SaTScan - Software for the spatial, temporal, and space-time scan statistics [Internet]. [cited 2024 Jan 22]. Available from: https://www.satscan.org/

28. Wickham H, François R, Henry L, Müller K, Vaughan D, Software P, et al. dplyr: A Grammar of Data Manipulation [Internet]. 2023 [cited 2024 Jan 22]. Available from: https://cran.r-project.org/web/packages/dplyr/index.html

29. Xie Y, Cheng J, Tan X, Allaire JJ, Girlich M, Ellis GF, et al. DT: A Wrapper of the JavaScript Library “DataTables” [Internet]. 2023 [cited 2024 Jan 22]. Available from: https://cran.r-project.org/web/packages/DT/index.html

30. Xu [aut H, cre, cph. findSVI: Calculate Social Vulnerability Index for Communities [Internet]. 2023 [cited 2024 Jan 30]. Available from: https://cran.r-project.org/web/packages/findSVI/index.html

31. Spinu V, Grolemund G, Wickham H, Vaughan D, Lyttle I, Costigan I, et al. lubridate: Make Dealing with Dates a Little Easier [Internet]. 2023 [cited 2024 Jan 22]. Available from: https://cran.r-project.org/web/packages/lubridate/index.html

32. Kleinman K, Hostovich S, Moosa A. rsatscan: Tools, Classes, and Methods for Interfacing with “SaTScan” Stand-Alone Software [Internet]. 2023 [cited 2024 Feb 13]. Available from: https://cran.r-project.org/web/packages/rsatscan/index.html

33. Pebesma E, Bivand R, Racine E, Sumner M, Cook I, Keitt T, et al. sf: Simple Features for R [Internet]. 2023 [cited 2024 Jan 22]. Available from: https://cran.r-project.org/web/packages/sf/index.html

34. Attali [aut D, cre. shinyjs: Easily Improve the User Experience of Your Shiny Apps in Seconds [Internet]. 2021 [cited 2024 Jan 30]. Available from: https://cran.r-project.org/web/packages/shinyjs/index.html

35. Chang W, RStudio, themes) TP (Bootswatch, font) LD (Lato, font) NW (News C, fonts) GC (Open S and R, et al. shinythemes: Themes for Shiny [Internet]. 2021 [cited 2024 Feb 6]. Available from: https://cran.r-project.org/web/packages/shinythemes/index.html

36. Wickham H, Software P, PBC. stringr: Simple, Consistent Wrappers for Common String Operations [Internet]. 2023 [cited 2024 Jan 22]. Available from: https://cran.r-project.org/web/packages/stringr/index.html

37. Walker K, Rudis B. tigris: Load Census TIGER/Line Shapefiles [Internet]. 2024 [cited 2024 Jul 8]. Available from: https://cran.r-project.org/web/packages/tigris/index.html

