## Supplementary Table 1 for "CalCORVID: A Dynamic RShiny Dashboard Approach to Visualize Spatiotemporal Clusters for Public Health Surveillance"

*Supplementary Table 1. Software and R packages used to develop the CalCORVID dashboard.*

| Name and version | Type | Description | Purpose/Relevance | Depends and Imports (required R packages) |
|---|---|---|---|---|
| R (4.0.4) | Software | Language and environment for statistical computing and graphics (17) | Programming language the dashboard is based off of | — |
| SaTScan (10.1) | Software | Free software that analyzes spatial, temporal, and spatiotemporal data using the space-time scan statistic (27) | User must obtain results from SaTScan to adapt dashboard code | — |
| dplyr (1.1.3) | R package | A fast, consistent tool for working with data frame like objects, both in and out of memory (28) | Manipulate and clean data to display on the dashboard | cli, generics, glue, lifecycle, magrittr, methods, pillar, R6, rlang, tibble, tidyselect, utils, vctrs |
| DT (0.18) | R package | Data objects in R can be rendered as HTML tables using the JavaScript library 'DataTables' (typically | Generate table to display cluster results on dashboard | htmltools, htmlwidgets, httpuv, jsonlite, magrittr, crosstalk, jquerylib, promises |

| Name and version | Type | Description | Purpose/Relevance | Depends and Imports (required R packages) |
|---|---|---|---|---|
| | | via R Markdown or Shiny) (29) | | |
| findSVI (0.1.2) | R package | Provided with year(s), region(s) and a geographic level of interest, 'findSVI' retrieves required variables from US census data and calculates SVI for communities in the specified area based on Centers for Disease Control/Agency for Toxic Substances and Disease Registry (CDC/ATSDR) SVI documentation (30) | Obtain Social Vulnerability Index (SVI) metrics displayed in cluster tooltip | cli, dplyr, magrittr, purrr, stringr, tidycensus, tidyr, tidyselect, rlang, utils |
| leaflet (2.0.4.1) | R package | Create and customize interactive maps using the ‘leaflet’ Javascript library and | Render map for overlaying cluster results in dashboard | crosstalk, htmltools, htmlwidgets, jquerylib, leaflet.providers, magrittr, methods, png, raster, |

| Name and version | Type | Description | Purpose/Relevance | Depends and Imports (required R packages) |
|---|---|---|---|---|
| | | 'htmlwidgets' package (24) | | RColorBrewer, scales, sp, stats, viridisLite, xfun |
| lubridate (1.7.10) | R package | Functions to work with date-times and time-spans, including parsing date-time data and algebraic manipulation of date-time and time-span objects (31) | Obtain dates at the beginning of a time frame (e.g., week start date) | methods, generics, timechange |
| rsatscan (1.0.7) | R package | Functions to write R data frames into SaTScan-readable formats, set SaTScan parameters, and run SaTScan software using a wrapper (32) | Run to obtain sample data | utils, foreign |
| sf (0.9.8) | R package | Support for simple features, a standardized way to | Read in the shapefile containing county boundaries to toggle as an | methods, classInt, DBI, graphics, grDevices, grid, magrittr, Rcpp, s2, stats, |

| Name and version | Type | Description | Purpose/Relevance | Depends and Imports (required R packages) |
|---|---|---|---|---|
| | | encode spatial vector data (33) | optional layer; calculate centroids for map zoom levels | tools, units, utils |
| shiny (1.7.1) | R package | A web application framework to build interactive web applications with R (18) | Underlying framework to develop RShiny dashboard | methods, utils, grDevices, httpuv, mime, jsonlite, xtable, fontawesome, htmltools, R6, sourcetools, later, promises, tools, crayon, rlang, fastmap, withr, commonmark, glue, bslib, cachem, lifecycle |
| shinyjs (2.1.0) | R package | Perform common useful JavaScript operations in Shiny apps that will greatly improve your apps without having to know any JavaScript (34) | Enable clicking images on the home page to switch tabs | digest, jsonlite, shiny |
| shinythemes (1.2.0) | R package | Themes for use with Shiny (35) | Set dashboard theme | shiny |

| Name and version | Type | Description | Purpose/Relevance | Depends and Imports (required R packages) |
|---|---|---|---|---|
| stringr (1.5.0) | R package | A consistent, simple and easy to use set of wrappers around the fantastic 'stringi' package (36) | Use for text string matching to identify CSV files | cli, glue, lifecycle, magrittr, rlang, stringi, vctrs |
| tigris | R package | Downloads TIGER/Line shapefiles from the United States Census Bureau and load into R as ‘sf’ objects (37) | Obtain county boundaries, state coordinates to orient leaflet map, and obtain polygons for clusters | stringr, magrittr, utils, rappdirs, httr, uuid, sf, dplyr, methods |
