## Supplementary Table 2 for "CalCORVID: A Dynamic RShiny Dashboard Approach to Visualize Spatiotemporal Clusters for Public Health Surveillance"

***Supplementary Table 2. Model specification used in SaTScan software to generate the sample display data. Default values were used for any unspecified parameters.***

| **[Setting] Parameter** | **Parameter Setting** |
|---|---|
| [Input] Study Period | September 21, 2021-February 8, 2022 |
| [Analysis] Analysis type | Prospective space-time |
| [Analysis] Probability Model | Discrete Poisson |
| [Analysis] Scan For areas With: | Low rates |
| [Analysis] Time Aggregation | Units: Day, Length: 7 |
| [Analysis→Spatial Window] Maximum Spatial Cluster Size | 50 km |
| [Analysis→Temporal Window] Maximum Temporal Cluster Size | 40 days |
| [Analysis→Temporal Window] Minimum Temporal Cluster Size | 1 day |
| [Analysis→Cluster Restrictions] Restrict low rate clusters to observed/expected less than or equal to: | 0.10 |
